# Acceleration of health deficit accumulation in late-life: Evidence of terminal decline in frailty index three years before death in the US Health and Retirement study

**DOI:** 10.1101/2020.10.11.20210732

**Authors:** Erwin Stolz, Hannes Mayerl, Emiel O. Hoogendijk, Joshua J. Armstrong, Regina Roller-Wirnsberger, Wolfgang Freidl

**Affiliations:** Institute of Social Medicine and Epidemiology, Medical University of Graz, Graz, Austria; Department of Epidemiology and Biostatistics, Amsterdam Public Health Research Institute, Amsterdam UMC - Location VU University Medical Center, Amsterdam, the Netherlands; Department of Health Sciences, Lakehead University, Thunder Bay, Canada; Department of Internal Medicine, Medical University of Graz, Graz, Austria

**Keywords:** frailty, geriatrics, death, aged, aged 80 and over, repeated rounds of survey

## Abstract

**Background:** Little is known about within-person frailty index (FI) changes during the last years of life. In this study, we assess whether there is a phase of accelerated health deficit accumulation (terminal health decline) in late-life.

**Material and methods:** 23,393 observations from up to the last 21 years of life of 5,713 deceased participants of the AHEAD cohort in the Health and Retirement Study were assessed. A FI with 32 health deficits was calculated for up to 10 successive biannual assessments (1995-2014), and FI changes according to time-to-death were analyzed with a piecewise linear mixed model with random change points.

**Results:** The average normal (pre-terminal) health deficit accumulation rate was 0.01 per year, which increased to 0.05 per year at approximately 3 years before death. Terminal decline began earlier in women and was steeper among men. The accelerated (terminal) rate of health deficit accumulation began at a FI value of 0.29 in the total sample, 0.27 for men, and 0.30 for women.

**Conclusion:** We found evidence for an observable terminal health decline in the FI following declining physiological reserves and failing repair mechanisms. Our results suggest a conceptually meaningful cut-off value for the continuous FI around 0.30.

## Introduction

Frailty has major implications for clinical practice and public health[1], and is defined as a vulnerability towards drastic health deterioration among older adults due to a cumulative decline in multiple physiological systems[2]. Specifically, it is thought that the gradual decline in physiologic reserves associated with normal aging accelerates in frailty, and consequently, homeostatic mechanisms start to fail. For frail older adults, even minor stressors can therefore result in disability[3], hospitalization[4] or death[5]. The established frailty index (FI)[6, 7], one of the most common and robust frailty assessment tools[8], measures an older person’s whole health based on the number of accumulated health deficits. For the FI, the number of present deficits a person has accumulated is divided by the total number of deficits considered (e.g. 6/30 = 0.20). In theory, the FI ranges from 0 (fit) to 1 (frail), but the empirical near-maximum (99th percentile) FI is consistently found below 0.70[9-11]. It is thought, that at this point, homeostasis reaches its limit, so that any additional health deficit cannot be sustained and would likely result in death. While this endpoint of the FI continuum is well established, it is less clear when frailty begins, that is, the tipping point[12] when homeostatic mechanisms start to fail and health deficit accumulation increases consequently. In other words, we are interested in *when* the pre-terminal phase characterized by stability or only minor health decline is replaced with the terminal phase of more rapid health decline which ends with death[13].

Currently, rather little is known about FI changes in late-life, and specifically during the last year(s) of life, that is, according to time-to-death (TTD), a time metric which better characterizes late-life health changes than time since birth (chronological age)[14]. The few available studies suggest, that the FI correlates more strongly with TTD than age[15], that there is likely a rapid FI-increase during the last year(s) of life[16,17], that individuals with higher FI scores accumulate further deficits more slowly compared to more robust older adults who catch-up during the very last years of life[18], and that men accumulate deficits near their time of death whereas this is extended over a longer period of time in women[19]. However, most of this evidence stems from cross-sectional analyses of a limited number or proportion of deceased older adults. To actually date and quantify the hypothesized acceleration in the rate of health deficit accumulation before death, non-linear within-person FI-changes during the last years of life should be assessed in a representative sample of deceased older adults for whom TTD is known. In this study, we perform such an analysis to answer whether there is an acceleration in the FI before death, how large it is, and when this typically occurs.

## Materials and methods

### Data

The oldest cohort (AHEAD) of the Health and Retirement Study (HRS)[20] provides population-representative longitudinal data for community-dwelling older adults in the United States and a near-complete mortality follow-up. AHEAD participants, who were born before 1924 were interviewed regarding their physical, functional, cognitive and psychological health biannually from 1993 onwards. Sufficient data for the calculation of the FI was available from 1995 (=baseline) to 2014, which resulted in a maximum of 10 repeated observations per person. When respondents were unwilling or unable to be (re-)interviewed, often due to medical or cognitive problems, a proxy respondent, usually a close family member, could provid information. Mortality follow-up for AHEAD participants based on the national mortality register lasted until August 2017 with a completion rate of 96.8%. During this period, 5846 (91.2%) of the AHEAD participants died with a known date of death, which constitutes the sample for this analysis.

### Variables

A frailty index (FI) was calculated from 32 identical health deficits in each wave (Supplementary Table 1) according to standard protocol[21]. The health deficits covered multiple physiological systems, and included chronic diseases, limitations in (instrumental) activities of daily living, mobility restrictions, symptoms, BMI-deficit, sensory impairments, and self-rated health and memory. All health deficits had less than 5% missing values, and the FI was calculated only for participants who provided valid information for at least 80% of the health deficits, which amounted to 97.3% (n=5,713) at baseline. Other variables were time to death (years), chronological age (years), age at death (years), and sex (male/female).

### Statistical Analysis

First, we calculated descriptive statistics to assess characteristics of FI values at baseline, follow-up waves, and at the last available measurement before death. Second, based on the pooled sample, we plotted non-linear FI trajectories by both chronological age and TTD using restricted cubic regression splines to obtain an overview of FI changes in late-life. Third, to analyze longitudinal within-person changes and to pinpoint when the normal (pre-terminal) health deficit accumulation rate is replaced by the hypothesized accelerated (terminal) health deficit accumulation rate, we applied a piecewise linear mixed model with random change points[22], a type of model often used to differentiate pre-terminal from terminal declines in cognitive functioning[23]. Specifically, this model takes the form of

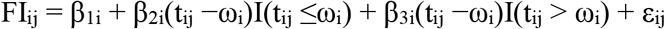

where observed FI measurements for the i^th^ person (i = 1, …, N) at time points t_ij_ (j = 1, …, n_i_) measured in years are modeled as a function of the individual-specific intercept (β_1i_), i.e. the expected FI value at the change point, the individual-specific normal (pre-terminal) FI slope (β_2i_) before the change point, the accelerated (terminal) individual-specific FI slope (β_3i_) after the change point, and the timing of the individual-specific change-point (ω_i_) itself. The individual-specific parameters consist of the population-average or fixed (β_10_,β_20_,β_30_, and ω_0_) effects and the random effects, that is, the individual-specific deviations from the population-level effects (u_1i_,u_2i_,u_3i_, and u_4i_) which account for both the heterogeneity between individuals and the autocorrelation between the varying number of repeated observations. Given the known sex-differences in the FI in both cross-sectional and longitudinal studies[24], we furthermore stratified all analyses by sex. All models were estimated with ‘brms’ (v.2.11.1), a front-end for ‘RStan’ (v.2.19.2) in R: A language and environment for statistical computing (v.4.0.2).

HRS data are freely available to researchers upon registration (https://hrs.isr.umich.edu/data-products), and the R-Markdown code reproducing all analyses and results is available online (https://osf.io/2vhzu/).

## Results

Median age at baseline was 78.9 (IQR=8.4) years, and 60.6% of the sample were women. Mean age at death was 89.0 (SD=6.2) years among women and 86.9 (SD=6.0) years among men. Median time of follow-up before death was 5.3 years (IQR=6.8, range=0-21.4) and the mean number of repeated assessments before death was 4.1 (SD=2.5, range=1-10). Specifically, 993 respondents (17.4%) provided only one FI assessment, 914 (16.0%) two, 853 (14.9%) three, 680 (11.9%) four, 606 (10.6%) five, 526 (9.2%) six, 472 (8.3%) seven, 286 (5.0%) eight, 220 (3.9%) nine, and 163 (2.9%) all ten assessments. In total, the 5,713 participants provided 23,393 observations. The median time between the last interview and time of death was 1.4 (IQR=1.2) years. 40.8% of the participants (n=2,329) were interviewed during their last 12 months, and 19.9% (n=1,136) even during the last six months of their life.

At baseline, mean/median FI values were 0.25/0.21 (SD=0.15/IQR=0.17) for men and 0.28/0.25 (SD=0.16/IQR=0.21) for women, and 95th/99th quantiles were 0.57/0.73 respectively 0.61/0.74. During follow-up, FI values increased with each subsequent assessment (Supplementary Figure 1). FI scores at baseline followed the characteristically right-skewed and sex-specific distribution typical of community-dwelling samples (plot A in Figure 1), whereas the distribution of FI values from the last available observation was more normal (plot B in Figure1), resembling clinical samples. FI values increased with both chronological age and TTD (plots C and D in Figure 1) but stronger and more progressively so with TTD (partial Pearson-r between FI and TTD adjusted for age=-0.29, p<0.001) than age (partial Pearson-r between FI and age adjusted for TTD=0.16, p<0.001). During follow-up, as sample size decreased, the proportion of proxy-interviews increased (e.g. 1995/1996=12.4%, 2004/2005=20.0%, 2014/2015=40.5%). Participants who provided fewer repeated observations were frailer on average at baseline (Supplementary Figure 2).

**Figure 1:**
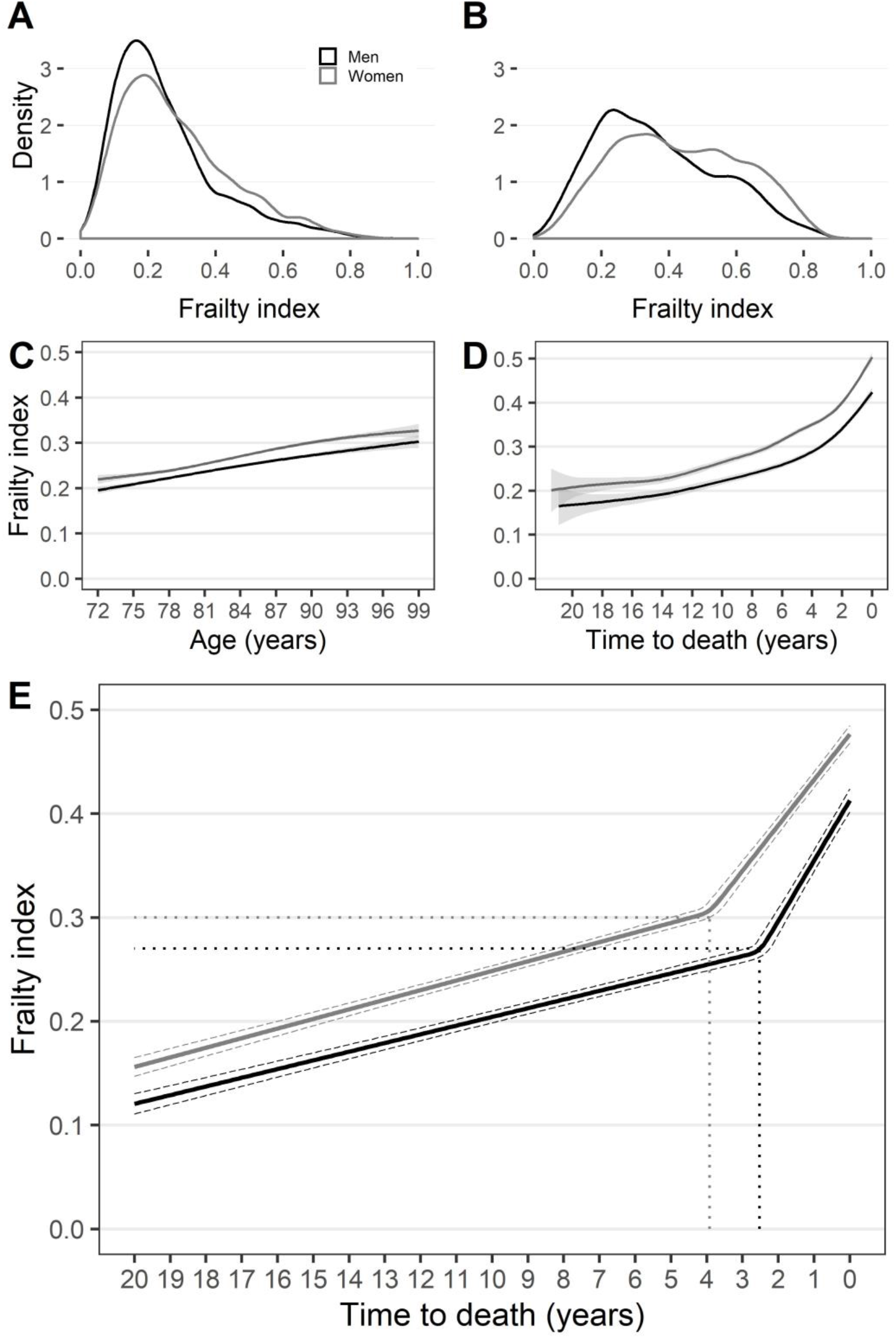
Frailty index distributions and trajectories for men and women. Plot A: Density plot of the distribution of frailty index values for men (black) and women (grey) at baseline; Plot B: Density plot of the distribution of frailty index values for men and women at the last available measurement per person; Plot C: Mean frailty index trajectories for men and women by age (restricted cubic regression spline) based on pooled data, shaded area refers to the 95% confidence interval; Plot D: Mean frailty index trajectories for men and women by time to death (restricted cubic regression spline) based on pooled data, shaded area refers to the 95% confidence interval; Plot E: Mean normal (pre-terminal) and accelerated (terminal) slope of frailty index based on piecewise linear mixed regression models for men and women. Solid lines refer to the estimated population-level (fixed) effects, dashed lines show 95% credible intervals.

The main results of this analysis, based on the piecewise linear mixed regression models, are shown in Table 1 and plot E in Figure 1. These show that the normal (pre-terminal) rate of health deficit accumulation of 0.01 per year increased five-fold (terminal decline) and that this change in pace occurred about 3 years before death. Terminal decline was steeper and occurred later in men compared to women. The average FI value at the onset of the accelerated health deficit accumulation phase was 0.29 (CI-95%=0.29-0.30) in total, and 0.27 (CI-95%=0.26-0.27) for men and 0.30 (CI-95%=0.30-0.31) for women. This means, women had both a longer terminal phase and were frailer during this period. Despite a steeper rate of health deficit accumulation in men after the change point, a gap between men and women remained at the end: estimated mean FI values right before death were 0.41 (CI-95%=0.40-0.42) among men and 0.48 (CI-95%=0.47-0.49) among women. However, due to the mean distance of 1.4 years before death in the assessments and the (bi-)linear nature of model, these are likely a down-biased estimates.

**Table 1:** Results of piecewise linear mixed regression model

|  | Total sample | Men | Women |
| --- | --- | --- | --- |
| | $\beta/u$ (CI-95%) | $\beta/u$ (CI-95%) | $\beta/u$ (CI-95%) |
| POPULATION-LEVEL EFFECTS |  |  |  |
| Intercept ($\beta_{10}$) | 0.29 (0.29, 0.30) | 0.27 (0.26, 0.27) | 0.30 (0.30, 0.31) |
| Normal (pre-terminal) FI slope ($\beta_{20}$) | 0.01 (0.01, 0.01) | 0.01 (0.01, 0.01) | 0.01 (0.01, 0.01) |
| Accelerated (terminal) FI slope ($\beta_{30}$) | 0.05 (0.05, 0.05) | 0.06 (0.05, 0.06) | 0.04 (0.04, 0.05) |
| Change point in years ($\omega_0$) | 3.26 (3.02, 3.49) | 2.52 (2.24, 2.81) | 3.92 (3.59, 4.26) |
| SD INDIVIDUAL-LEVEL EFFECTS |  |  |  |
| Intercept ($u_{i1}$) | 0.11 (0.11, 0.12) | 0.10 (0.10, 0.11) | 0.12 (0.11, 0.12) |
| Normal (pre-terminal) FI slope ($u_{i2}$) | 0.00 (0.00, 0.00) | 0.00 (0.00, 0.00) | 0.00 (0.00, 0.01) |
| Accelerated (terminal) FI slope ($u_{i3}$) | 0.04 (0.04, 0.04) | 0.05 (0.04, 0.05) | 0.03 (0.03, 0.04) |
| Change point ($u_{i4}$) | 0.80 (0.75, 0.85) | 0.89 (0.80, 0.98) | 0.72 (0.66, 0.78) |
| MODEL INFORMATION |  |  |  |
| Number of observations | 23,484 | 8,697 | 14,787 |
| Number of respondents | 5,713 | 2,252 | 3,461 |
| R-squared | 0.17 (0.17, 0.19) | 0.16 (0.14, 0.17) | 0.20 (0.19, 0.21) |

Weakly informative priors (student (0,10)) were used and estimation method was Hamiltonian Monte Carlo (HMC) with no-U-turn (NUTS) sampling, 6 chains with each 5,000 iterations and 1,000 iterations warm-up. All $\hat{r}$ values = 1.00, and all effective sample sizes > 500. R-squared refers to population-level (fixed) effects, 95%-CI = 95% credible interval.

## Discussion

In this study, we analyzed whether, how, and at which point before death the rate of health deficit accumulation in the FI accelerates. Frailty is thought to result from an accelerated decline in available physiologic reserves as these are increasingly required to compensate for changes associated with normal aging[24]. In consequence, repair mechanisms start to fail due to exceeding recovery time and damaged recovery processes[7], which leads to ever more unrepaired deficits. In this paper, we argue that failing repair mechanism should manifest soon in an empirically detectable acceleration of the rate of health deficit accumulation in late-life. To uncover this change in pace, we partitioned the observed FI trajectories towards death in a large sample of deceased participants with the help of a statistical model into a normal (or pre-terminal) and an accelerated (or terminal) rate of health deficit accumulation. The central conclusion that can be drawn from our results is that the health deficit accumulation rate indeed accelerates substantially about 3 years before death and that the average change or tipping point[12] in the FI that separates normal ageing from terminal decline was close to 0.30 in our study.

We interpret this within-person change in pace of late-life health deficit accumulation as the transition point into or the onset of frailty within the inherently continuous FI[6]. This interpretation is compatible with the verbal descriptors “frail” of the last three categories of the Clinical Frailty Scale (CFS), which begins with “mildly frail” and an associated mean FI of 0.27[26]. Our results can be tied back to the issue of a cut-off value for the FI. The question whether an old person is frail or not (yes/no) is important for both clinical practice, e.g. screening for frailty to prognosticate risk and balance between benefits and harms of interventions, and public health, e.g. with regard to frailty prevalence[1]. Therefore, various cut-offs for the presence of frailty in the FI have been suggested, e.g. 0.20[21], 0.25[27], or 0.35[28], although these round numbered cut-offs have little conceptual or empirical grounding. Our results suggest a cut-off close to 0.30, which is higher than the most often referenced FI-cut-off of 0.25, and (slight) differences between men and women. Hoogendijk et al.[29] recently also suggested a higher cut-off for women than men based on the ability to predict mortality. The FI cut-off values in that study, however, were lower compared to the present study, which may be due to country and/or methodological differences, particularly the fact that we assessed the FI multiple times within the same individuals and included only deceased participants.

The results of our study also relate to findings from previous research regarding FI dynamics in late-life. We found that women had consistently higher FI values throughout their last years of life, and that the phase of accelerated health deficit accumulation was a more drawn-out process among them compared to men who accumulated health deficits faster during the very last years of life[19], which is in line with the known male-female health-survival paradox[24]. Also, we were able to confirm a higher correlation of the FI with TTD compared to age[15], and that there is indeed a substantial increase in the rate of deficit accumulation in late-life – a five-fold increase amounting to two additional deficits per year in the terminal phase – as previously assumed[16]. Our results also relate to the question as to when the end-of-life phase begins[17]. According to a recent systematic review[30], this phase begins with the onset of terminal decline, which, according to our results based on a measure of a person’s whole health (the FI), would be around 3 years before death. These results are compatible with findings from studies on the onset of terminal decline in cognition[31] and motor function[32]. To the best of our knowledge, our study is the first to comprehensively assess changes in the FI in relation to TTD – including the estimation of the timing of accelerated health deficit accumulation – which adds to the existing literature both regarding FI dynamics and regarding terminal decline[13] more generally.

In our analysis, the sub-maximum FI-limit (99% percentile) was slightly above 0.70 and thus higher than in most studies[9-11]. This is likely due to the fact, (1) that we used a sample of deceased older adults of advanced age (median age at baseline was 79 years), (2) that we did not exclude participants who dropped out of the study during follow-up as mixed models make use of all available observations, and (3) that we included participants whose FIs were calculated based on information from proxy-interviews when the original participant would or could not participate due to severely impaired physical and/or cognitive health. Indeed, if based on self-reported data alone, FI scores were lower on average and the sub-maximum limit at baseline was 0.65 instead of 0.74. Proxy-interviews from close family members and caregivers, however, can be a reliable alternative source of information[33] in case of compromised health among the oldest old, and both increase statistical power and reduce selection bias[34], which is particularly important for the study of health changes in late-life. Future studies should assess the reliability and impact of including proxy-interviews with regard to the FI more thoroughly, for example regarding FI trajectories and the relationship with adverse health outcomes.

In this study, we relied on the same methodological framework that is used for the assessment of terminal decline in cognitive functioning[23], i.e. we tracked FI changes over TTD postdictively among deceased older adults. Since, the date of death is not known in advance, however, the information provided in this study of an accelerated health deficit accumulation rate in the very last years before death is of limited direct clinical relevance. Nonetheless, the FI cut-off values suggested by our study, and the characteristics of the accelerated rate of health deficit accumulation add to the understanding of late-life frailty dynamics, and could also be of predictive use. Future studies based on routinely collected data in clinical practice could, for example, assess steep, ‘terminal-like’ acceleration in FI growth as an indicator that a critical tipping point[12] in a patient’s health has been reached.

The current study has several strengths. First, this is the first study to analyze the timing of the onset and the size of terminal health decline in the FI based on extensive longitudinal data. Second, we provided a conceptually meaningful empirical estimate of a cut-off value for the FI. Third, our analysis was based on a considerable number of repeated FI observations from decedents of a large, population-representative cohort with near-complete mortality. This is important because TTD – which better characterizes late-life health changes than age[14] – is known only among decedents and a low proportion of deceased sample participants after only a few years of follow-up would be selective and likely lead to biased FI estimates.

However, there are also several noteworthy limitations to this study. First, it is unclear whether the results based on the oldest cohort in HRS – all participants had to reach at least age 72 to be included – generalize to later birth cohorts and other geographic contexts. Previous research has indicated considerable cross-national differences in FI dynamics[36] as well as increased FI values among later birth cohorts[36] which, together with varying life-expectancy may lead to different results. Thus, our calculations should be repeated with other long-running, population-representative, cohort studies with near complete mortality such as the Longitudinal Aging Study Amsterdam[38], or the Honolulu-Asia Aging Study[11]. Second, due to the inclusion of proxy-respondents, we could not include any performance-based or psychological health deficits, which may lead to an underestimation of frailty levels[39]. Third, despite the comparatively high retention rate and the considerable proportion of proxy-interviews in HRS compared to other studies, its biannual cycle and the high median age at baseline resulted in relatively few available repeated observations per person (around four on average). This forestalled, for example, estimating the correlation coefficients between individual-level (random) effects – and to assess whether frailer older adults have less steep terminal acceleration rates and vice-versa as suggested by Rockwood et al.[18] – since the model would not converge upon adding six additional individual-effect parameters. Fourth, the non-trivial temporal distance between the last FI assessment and death limits our ability to reliably assess FI-changes in the very last year of life. The last two limitations may be remedied only with more intensive population-level data (e.g. 10+ repeated quarterly assessments during the last years of life), which might become available in the near future from routinely collected primary care electronic health records[17,35].

## Conclusion

To conclude, this study conducted in a large cohort of deceased older adults indicates that the rate of health deficit accumulation accelerates substantially in the last three years of life before death, which we interpret as the consequence of steeply declining physiologic reserves and the onset of failing repair mechanisms. According to our results, acceleration of health deficit accumulation begins on average at an FI value close to 0.30, which we thus suggest as a conceptually meaningful and empirically derived cut-off value for the presence of frailty in the continuous FI.

## Data Availability

HRS data are freely available to researchers upon registration, and the R-Markdown code reproducing all analyses and results is available online (OSF).

https://hrs.isr.umich.edu/data-products

https://osf.io/2vhzu/

---

**Supplementary Table 1:** Health deficits of frailty index

| Health deficit | Cut-point(s) |
| --- | --- |
| 1. Doctor told you had heart problems | no = 0, yes = 1 |
| 2. Doctor told you had high blood pressure | no = 0, yes = 1 |
| 3. Doctor told you had a stroke | no = 0, yes = 1 |
| 4. Doctor told you had diabetes | no = 0, yes = 1 |
| 5. Doctor told you had cancer | no = 0, yes = 1 |
| 6. Doctor told you had lung disease | no = 0, yes = 1 |
| 7. Doctor told you had arthritis | no = 0, yes = 1 |
| 8. Doctor told you had mental health problem(s) | no = 0, yes = 1 |
| 9. Difficulty dressing | no = 0, yes = 1 |
| 10. Difficulty walking across room | no = 0, yes = 1 |
| 11. Difficulty using toilet | no = 0, yes = 1 |
| 12. Difficulty bathing/showering | no = 0, yes = 1 |
| 13. Difficulty eating | no = 0, yes = 1 |
| 14. Difficulty getting out of bed | no = 0, yes = 1 |
| 15. Difficulty making phone calls | no = 0, yes = 1 |
| 16. Difficulty managing money | no = 0, yes = 1 |
| 17. Difficulty walking one block | no = 0, yes = 1 |
| 18. Difficulty getting up from chair | no = 0, yes = 1 |
| 19. Difficulty sitting (2 hours) | no = 0, yes = 1 |
| 20. Difficulty stooping/kneeling/crouching | no = 0, yes = 1 |
| 21. Difficulty raising arms | no = 0, yes = 1 |
| 22. Difficulty picking coin from table | no = 0, yes = 1 |
| 23. Self-rated health | excellent = 0, very good = 0.25, good = 0.5, fair = 0.75, poor = 1 |
| 24. Self-rated memory | excellent = 0, very good = 0.25, good = 0.5, fair = 0.75, poor = 1 |
| 25. Self-rated eyesight close | excellent = 0, very good = 0.25, good = 0.5, fair = 0.75, poor = 1 |
| 26. Self-rated eyesight distance | excellent = 0, very good = 0.25, good = 0.5, fair = 0.75, poor = 1 |
| 27. Self-rated hearing | excellent = 0, very good = 0.25, good = 0.5, fair = 0.75, poor = 1 |
| 28. BMI-deficit | BMI ≥ 18.5 and BMI $<$ 25 = 0, BMI ≥ 25 and BMI ≤ 30 = 0.5, BMI $>$ 18.5 or BMI $>$ 30 = 1 |
| 29. Fallen down in last two years | no = 0, yes = 1 |
| 30. Often troubled by pain | no = 0, yes = 1 |
| 31. Incontinence | no = 0, yes = 1 |
| 32. Broken hip | no = 0, yes = 1 |

**Supplementary Figure 1:**
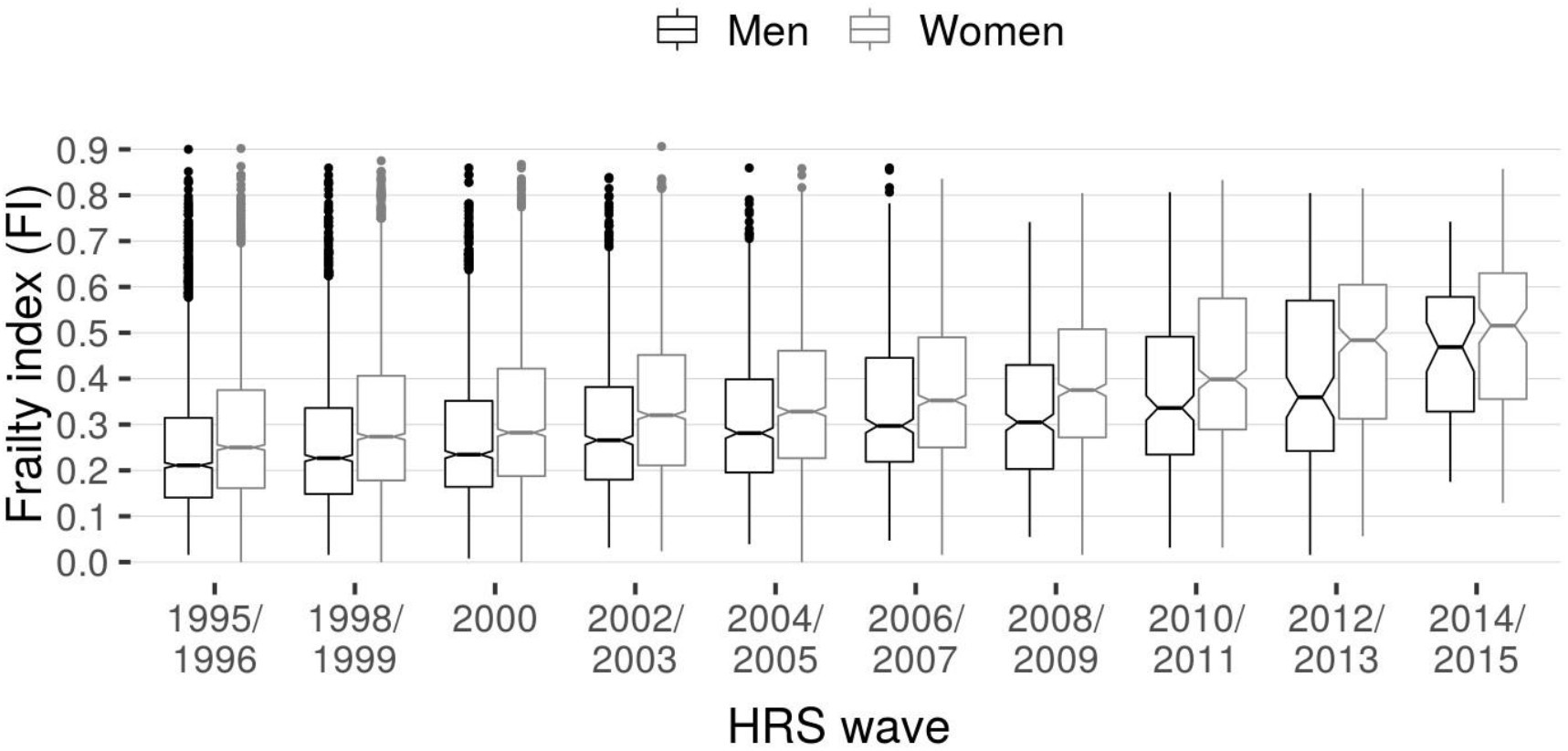
Frailty index by HRS wave. Black boxplots refer to men, grey boxplots refer to women. The boxes represent the interquartile range (IQR, covering data from the 25th-75th percentile), the whiskers extend 1.5*IQR from the hinges. Single dots refer to outlying observations beyond the whiskers. Boxes are segmented by the median and the notches around the median indicate its 95% confidence interval. Note that sample size decreased with each wave: n1995/1996=5,713, n1998/1999=4,598, n2000=3,737, n2002/2003=2,925, n_2004_/2005=2,280, n_2006_/2007=1,695, n_2008_/2009=1,201, n_2010_/2011=706, n_2012_/2013=438, and n2014/2015=191.

**Supplementary Figure 2:**
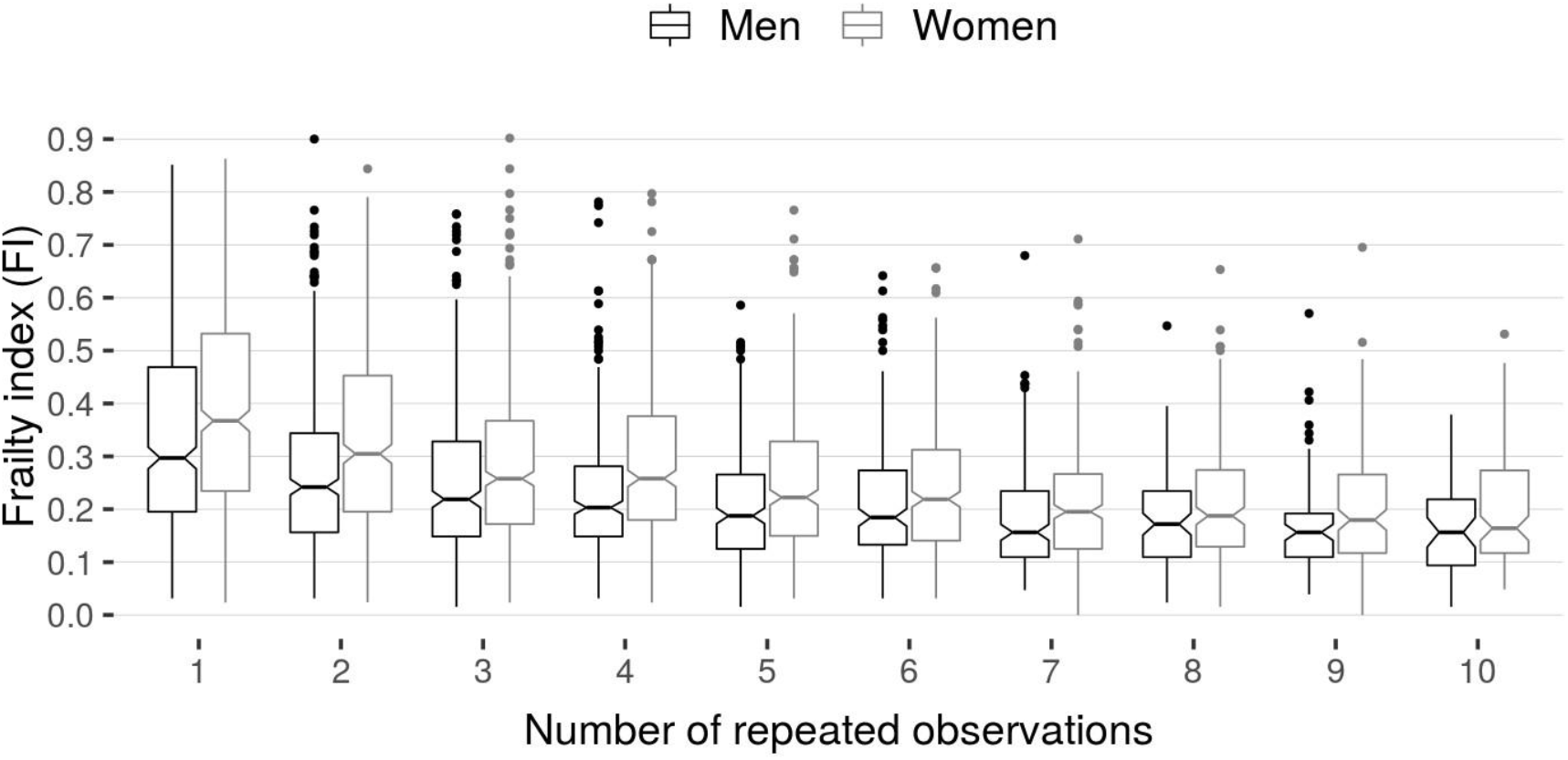
Frailty index values at baseline by number of repeated observations available per person. Black boxplots refer to men, grey boxplots refer to women. The boxes represent the interquartile range (IQR, covering data from the 25th-75th percentile), the whiskers extend 1.5*IQR from the hinges. Single dots refer to outlying observations beyond the whiskers. Boxes are segmented by the median and the notches around the median indicate its 95% confidence interval.

## References

1. Hoogendijk, EO, Afilalo, J, Ensrud, KE, Kowal, P, Onder, G, and Fried, LP. Frailty: implications for clinical practice and public health. Lancet 2019;394:1365–1375. https://doi.org/10.1016/S0140-6736(19)31786-6.

2. Clegg, A, Young, J, Iliffe, S, Rikkert, MO, and Rockwood, K. Frailty in elderly people. Lancet 2013;381:752–762. https://doi.org/10.1016/S0140-6736(12)62167-9.

3. Kojima, G. Frailty as a predictor of disabilities among community-dwelling older people: a systematic review and meta-analysis. Disability and Rehabilitation 2017;39:1897–1908. https://doi.org/10.1080/09638288.2016.1212282.

4. Kojima, G. Frailty as a predictor of hospitalisation among community-dwelling older people: a systematic review and meta-analysis. J Epidemiol Community Health 2016;70:722–729. https://doi.org/10.1136/jech-2015-206978.

5. Kojima, G, Iliffe, S, and Walters, K. Frailty index as a predictor of mortality: a systematic review and meta-analysis. Age Ageing 2018;47:193–200. https://doi.org/10.1093/ageing/afx162.

6. Rockwood, K and Mitnitski, A. Frailty in relation to the accumulation of deficits. J Gerontol A Biol Sci Med Sci. 2007;62:722–727. https://doi.org/10.1093/gerona/62.7.722.

7. Rockwood, K and Howlett, SE. Age-related deficit accumulation and the diseases of ageing. Mech Age Dev 2019;180:107–116. https://doi.org/10.1016/j.mad.2019.04.005.

8. Dent, E, Kowal, P, and Hoogendijk, EO. Frailty measurement in research and clinical practice: A review. Eur J Intern Med 2016;31:3–10. https://doi.org/10.1016/j.ejim.2016.03.007.

9. Rockwood, K and Mitnitski, A. Limits to deficit accumulation in elderly people. Mech Age Dev 2006;127:494–496. https://doi.org/10.1016/j.mad.2006.01.002.

10. Bennett, S, Song, X, Mitnitski, A, and Rockwood, K. A limit to frailty in very old, community-dwelling people: a secondary analysis of the Chinese longitudinal health and longevity study. Age Ageing 2013;42:372–377. https://doi.org/10.1093/ageing/afs180.

11. Armstrong, JJ, Mitnitski, A, Launer, LJ, White, LR, and Rockwood, K. Frailty in the Honolulu-Asia Aging Study: Deficit Accumulation in a Male Cohort Followed to 90% Mortality. J Gerontol A Biol Sci Med Sci 2015;70:125–131. https://doi.org/10.1093/gerona/glu089.

12. Rikkert, MGM, Melis RJF. Rerouting geriatric medicine by complementing static frailty measures with dynamic resilience indicators ofl recovery potential. Front Phys 2019;10:723. https://doi.org/10.3389/fphys.2019.00723.

13. Gerstorf, D and Ram, N. Inquiry Into Terminal Decline: Five Objectives for Future Study. Gerontologist 2013;53:727–737. https://doi.org/10.1093/geront/gnt046.

14. Gerstorf, D, Ram, N, Lindenberger, U, and Smith, J. Age and time-to-death trajectories of change in indicators of cognitive, sensory, physical, health, social, and self-related functions. Dev Psychol 2013;49:1805–1821. https://doi.org/10.1037/a0031340.

15. Kulminski, A, Yashin, A, Arbeev, K, et al. Cumulative index of health disorders as an indicator of the aging-associated proccesses in elderly: Results From Analyses of the National Long Term Care Survey. Mech Ageing Dev 2007;128:250–258. https://doi.org/10.1016/j.mad.2006.12.004.

16. Rockwood, K and Mitnitski, A. Frailty defined by deficit accumulation and geriatric medicine defined by frailty. Clin Geriatr Med 2011;27:17–26. https://doi.org/10.1016/j.cger.2010.08.008.

17. Stow, D, Matthews, FE, and Hanratty, B. Frailty trajectories to identify end of life: a longitudinal population-based study. BMC Med 2018;16:171. https://doi.org/10.1186/s12916-018-1148-x.

18. Rockwood, K, Rockwood, MRH, and Mitnitski, A. Physiological Redundancy in Older Adults in Relation to the Change with Age in the Slope of a Frailty Index. J Am Geriatr Soc 2010;58:318–323. https://doi.org/10.1111/j.1532-5415.2009.02667.x.

19. Kulminski, A, Ukraintseva, SV, Akushevich, I, Arbeev, KG, Land, K, and Yashin, AI. Accelerated accumulation of health deficits as a characteristic of aging. Exp Gerontol 2007;42:963–970. https://doi.org/10.1016/j.exger.2007.05.009.

20. Sonnega A, Fraul JD, Ofstedal MB, Langa KM, Phililips JWR, Weir DR. Cohort profile: the Health and Retirement Study (HRS). Int J Epidemiol 2014;43:576-585. DOI: 10.1093/ije/dyu067.

21. Searle, SD, Mitnitski, A, Gahbauer, EA, Gill, TM, and Rockwood, K. A standard procedure for creating a frailty index. BMC Geriatrics 2008;8:24. https://doi.org/10.1186/1471-2318-8-24.

22. Brilleman, SL, Howe, LD, Wolfe, R, and Tilling, K. Bayesian Piecewise Linear Mixed Models With a Random Change Point: An Application to BMI Rebound in Childhood. Epidemiology 2017;28:827–833. https://doi.org/10.1097/EDE.0000000000000723.

23. Karr, JE, Graham, RB, Hofer, SM, and Muniz-Terrera, G. When does cognitive decline begin? A systematic review of change point studies on accelerated decline in cognitive and neurological outcomes preceding mild cognitive impairment, dementia, and death. Psychol Aging 2018;33:195–218. https://doi.org/10.1037/pag0000236.

24. Gordon, EH, Peel, NM, Samanta, M, Theou, O, Howlett, SE, and Hubbard, RE. Sex differences in frailty: A systematic review and meta-analysis. Exp Geront 2017;89:30–40. https://doi.org/10.1016/j.exger.2016.12.021.

25. Taffett, GE. Physiology of Aging. In: Geriatric Medicine: An Evidence-Based Approach. Ed. by Cassel, CK, Leipzig, RM, Cohen, HJ, Larson, EB, Meier, DE, and Capello, CF. New York, NY: Springer, 2003:27–35. https://doi.org/10.1007/0-387-22621-4_3.

26. Rockwood K, Song X, MacKnight C, Bergman H, Hogan DB, McDowell I, Mitnitski A. A global clinical measure of fitness and frailty in elderly people. Can Med Ass J 2005;30:489–495. https://doi.org/10.1053/cmaj.050051.

27. Song, X, Mitnitski, A, and Rockwood, K. Prevalence and 10-Year Outcomes of Frailty in Older Adults in Relation to Deficit Accumulation. J Am Geriatr Soc 2010;58:681–687. https://doi.org/10.1111/j.1532-5415.2010.02764.x.

28. Kulminski, AM, Ukraintseva, SV, Kulminskaya, IV, Arbeev, KG, Land, K, and Yashin, AI. Cumulative deficits better characterize susceptibility to death in elderly people than phenotypic frailty: lessons from the Cardiovascular Health Study. J Am Geriatr Soc 2008;56:898–903. https://doi.org/10.1111/j.1532-5415.2008.01656.x.

29. Hoogendijk, EO, Stenholm, S, Ferrucci, L, Bandinelli, S, Inzitari, M, and Cesari, M. Operationalization of a frailty index among older adults in the InCHIANTI study: predictive ability for all-cause and cardiovascular disease mortality. Aging Clin Exp Res 2020;32:1025–1034. https://doi.org/10.1007/s40520-020-01478-3.

30. Cohen-Mansfield, J, Skornick-Bouchbinder, M, and Brill, S. Trajectories of End of Life: A Systematic Review. J Gerontol B Psychol Sci Soc Sci 2018;73:564–572. https://doi.org/10.1093/geronb/gbx093.

31. Wilson, RS, Beck, TL, Bienias, JL, and Bennett, DA. Terminal cognitive decline: accelerated loss of cognition in the last years of life. Psychosom Med 2007;69:131–137. https://doi.org/10.1097/PSY.0b013e31803130ae.

32. Wilson, RS, Segawa, E, Buchman, AS, Boyle, PA, Hizel, LP, and Bennett, DA. Terminal Decline in Motor Function. Psychol Aging 2012;27:998–1007. https://doi.org/10.1037/a0028182.

33. Ostbye, T, Tyas, S, Mcdowell, I, and Koval, J. Reported activities of daily living: agreement between elderly subjects with and without dementia and their caregivers. Age Ageing 1997;26:99–106. https://doi.org/10.1093/ageing/26.2.99.

34. Kelfve, S, Thorslund, M, and Lennartsson, C. Sampling and non-response bias on health-outcomes in surveys of the oldest old. Eur J Ageing 2013;10:237–245. https://doi.org/10.1007/s10433-013-0275-7.

35. Clegg, A, Bates, C, Young, J, et al. Development and validation of an electronic frailty index using routine primary care electronic health record data. Age Ageing 2016;45:353–360. https://doi.org/10.1093/ageing/afw039.

36. Stolz, E, Mayerl, H, and Freidl, W. Fluctuations in frailty among older adults. Age Ageing 2019;48:547–552. https://doi.org/10.1093/ageing/afz040.

37. Yu, R, Wong, M, Chong, KC, et al. Trajectories of frailty among Chinese older people in Hong Kong between 2001 and 2012: an age-period-cohort analysis. Age Ageing 2018;47:254–261. https://doi.org/10.1093/ageing/afx170.

38. Hoogendijk, EO, Theou, O, Rockwood, K, Onwuteaka-Philipsen, BD, Deeg, DJH, and Huisman, M. Development and validation of a frailty index in the Longitudinal Aging Study Amsterdam. Aging Clin Exp Res 2017;29:927–933. https://doi.org/10.1007/s40520016-0689-0.

39. Theou O, O’Connel MDL, King-Kallimanis BL, O’Halloran AM, Rockwood K, Kenny RA. Measuring frailty using self-report and test-based health measures. Age Ageing 2015;44:471–477.

